# Children’s emotional wellbeing during spring 2020 COVID-19 restrictions: a qualitative study with parents of young children in England

**DOI:** 10.1101/2021.07.02.21259900

**Authors:** Stephanie Chambers, Joanne Clarke, Ruth Kipping, Rebecca Langford, Rachel Brophy, Kim Hannam, Hilary Taylor, Kate Willis, Sharon A Simpson

**Author notes:** **Funding statement** This work was funded by the NIHR School for Public Health Research and NIHR funding for the NAP SACC UK trial (2019-3426). **Author contributions** The study was conceived by JC, RK, SC, KW, HT, RB, KH, SS and RL. JC led the study with oversight from RL and RK. Interviews were conducted by JC, SC and KW. Coding of the data was performed by JC, HT and RB. SC produced the first draft of the manuscript, with all other authors providing critical review and intellectual content. All authors read and approved the final manuscript. **Data availability statement** Anonymised study data will be made available under controlled access via the University of Bristol data repository. Requests for controlled data will be referred to an appropriate Data Access Committee for approval before data can be shared with bona fide researchers, after their host institution has signed a Data Access Agreement. **Conflict of interest disclosure** The authors declare they have no conflicts of interest. **Ethics approval statement** Ethical approval for the study was granted by University of Bristol Faculty of Health Sciences Research Ethics Committee (Ref: 106002). Participant consent was recorded on a separate audio file before the interview commenced.

## Abstract

**Background:** During COVID-19 restrictions in England in spring 2020, early years settings for young children were closed to all but a small percentage of families, social contact was limited and play areas in parks were closed. Concerns were raised about the impact of these restrictions on young children’s emotional wellbeing. The aim of this study was to explore parents’ perceptions of young children’s emotional wellbeing during these COVID-19 restrictions.

**Methods:** We interviewed 20 parents of children 3-4 years due to begin school in England in September 2020. Interviews were conducted via telephone (n=18) and video call (n=2), audio-recorded and transcribed verbatim. Interviews focused on childcare arrangements, children’s behaviour and transition to school. A sample of transcripts were coded line-by-line to create a coding framework, which was subsequently applied to the remaining transcripts. Coded data were then analysed using a nurture lens to develop themes and further understanding.

**Results:** Participants were predominantly mothers (n=16), White British (n=10), and educated to degree level (n=13), with half the sample living in the highest deprivation quintile in England (n=10). Five were single parents. Three themes developed from nurturing concepts were identified: creating age-appropriate explanations; understanding children’s behaviour; concerns about school transition. Parents recognised their children’s emotional wellbeing was impacted but attempted to support their young children whilst looking ahead to their transition to primary school.

**Conclusions:** This study is one of the first to examine in-depth the impact of COVID-19 restrictions on young children’s emotional wellbeing. The longer-term impacts are not yet understood. Although young children may be unable to understand in detail what the virus is, they undoubtedly experience the disruption it brings to their lives. The wellbeing of families and children needs to be nurtured as they recover from the effects of the pandemic to allow them to thrive.

**Key messages:** COVID-19 restrictions are predicted to have a negative impact on young children.

We interviewed parents of children in England due to begin primary school to understand their experiences of COVID-19 restrictions.

Nurture concepts helped us understand the challenges families faced.

Key themes identified were: creating age-appropriate explanations; understanding children’s behaviour; and concerns about school transition.

We suggest a nurturing approach to recovery to best support children and their families.

## Introduction

The COVID-19 pandemic has been an unprecedented event for all, including families with young children. In England between March – May 2020, early years childcare settings, hereafter called nurseries, were closed to all except children of essential workers and vulnerable children. There were limits on leaving one’s home and accessing play areas in parks, whilst non-essential shops were closed and social contact between households forbidden. Nine million jobs were furloughed with the UK Government paying 80% of the salary of employees unable to work (HM Revenue and Customs, 2020).

Nurseries provide a safe place for children to develop and thrive. During COVID-19 restrictions in Spring 2020, only one in 20 vulnerable children in England were attending childcare/schooling, despite being eligible to attend (Department for Education, 2020). In England, more children returned to nurseries in June 2020, although often for fewer days or hours than before. Concerns were raised about the impact of the pandemic on children’s health and wellbeing. Interim results from an ongoing study found that teachers were concerned about new school starters’ communication, language development and literacy (Bowyer-Crane et al., 2021). There were also concerns about young children’s social and emotional wellbeing (The Children’s Society, 2020). Pre-school child development includes learning to play with others, cooperate and develop attachments to secondary caregivers. The effect of high parental stress on family life and parenting while educational and community settings were closed is of particular concern (Benner & Mistry, 2020; Griffith, 2020; Masten & Motti-Stefanidi, 2020; Yoshikawa et al., 2020). Previous studies have shown that parents experiencing greater stress levels are less likely to engage in responsive care with their children. Responsive care is important for children’s development, and is likely to be even more essential during disruptions (Yoshikawa et al., 2020).

Researchers have hypothesised the likely impact of COVID-19 restrictions on young children based on theory and research from previous emergency situations (Benner & Mistry, 2020; Brown, Doom, Lechuga-Peña, Watamura, & Koppels, 2020; Masten & Motti-Stefanidi, 2020; Prime, Wade, & Browne, 2020; Yoshikawa et al., 2020). Most studies from the COVID-19 pandemic focus on older age groups (Royal College of Paediatrics and Child Health C& YPE team, 2021). There are some quantitative findings emerging, for example, parents in the Co-SPYCE study were concerned about the impact of the pandemic on pre-school children’s wellbeing and wanted greater support in managing children’s behaviours and emotions (Dodd, Westbrook, & Lawrence, 2020). Public Health Scotland surveyed parents of children aged 2-7 years with half reporting worse behaviour and mood in their children during COVID-19 restrictions (Public Health Scotland, 2020). Paul et al. (2021) found a greater persistence of reported emotional and behavioural difficulties in children after age two during the pandemic compared with pre-pandemic reports.

These studies provide an important overview but do not provide in-depth understanding of the issues faced by families and the techniques families use to support their pre-school children through the pandemic disruption. This study explored parents’ perceptions of their pre-school children’s emotional wellbeing during COVID-19 restrictions. The impact of COVID-19 on children’s social and emotional development will likely be felt by families in the long-term. It is essential we understand families’ experiences of this time to support them appropriately, and to inform future responses to similar emergency situations.

### Nurturing

We use nurturing as a lens to examine parenting during COVID-19 restrictions. Nurture is a recurring theme in the literature on how families might manage their children’s wellbeing during the pandemic (Brown et al., 2020; Masten & Motti-Stefanidi, 2020; Prime et al., 2020; Walsh, 2020; Yoshikawa et al., 2020). Nurture has its basis in attachment theory (Bowlby, 1958), which considers the relationship between a child and their primary caregiver. Where children have attached securely to their primary caregiver, there is warmth and responsiveness (Ainsworth, Blehar, Waters, & Wall, 2015). From the strength of this relationship, the child can develop and explore their environment with a feeling of security. However, where this relationship has not been nurtured, the child is more likely to experience social and emotional difficulties, and is less likely to form appropriate secondary attachments to further their development and enable them to flourish. Evidence from the literature on emergency situations has consistently stressed the importance of close relationships in supporting children through adversity (Masten & Motti-Stefanidi, 2020).

## Method

### Research Design

The study aim emerged from a wider qualitative study exploring the impact of COVID-19 restrictions on the health, wellbeing and behaviours of children three-four years due to begin primary school in September 2020. Impact on eating and activity is reported elsewhere (Clarke et al., 2020). As emotional wellbeing is so important at this age, we sought to analyse these data separately using a nurturing framework to help make sense of the data and identify recommendations.

### Recruitment

We recruited parents of pre-school children from nurseries in the West Midlands and South West of England participating in a trial led by this study’s authors (https://napsaccuk.blogs.bristol.ac.uk/). Nurseries emailed parents to inform them about the study. We also publicised the study via social media sites in these areas. Parents registered their interest via a website and provided demographic information and postcode data to derive deprivation level (https://imd-by-postcode.opendatacommunities.org/imd/2019) to enable sampling for diversity. We sampled participants from a pool of 85 who had expressed their interest in participating. We emailed these parents an information sheet and consent form, and telephoned to confirm participation and arrange a suitable interview time. JC, KW and SC conducted 20 interviews via video call (n = 2) or by telephone (n = 18) after gaining verbal consent. Interviews were audio-recorded and transcribed verbatim. An overview of the topic guide can be found in Table 1. Participants received a £30 shopping voucher following interview.

**Table 1.** Topic guide

| Domain | Questions |
| --- | --- |
| **Emotional wellbeing** | How has your child been feeling about the coronavirus restrictions?<br>What effects (if any) do you think the restrictions have had on your child's well-being?<br>Have the restrictions had any effect on your child's mood and feelings? (positive or negative)<br>Have you noticed any changes at all in your child's behaviour? (positive or negative)<br>What is the impact on the child of any reduced social contact with other children, friends and family? |
| **Transition to school** | How are you feeling about your child starting school in September?<br>How is your child feeling?<br>Any thoughts about whether child will start in September, or whether you may delay starting school?<br>Are there any things you would have been doing to help prepare the child for starting school that you aren't able to do because of the disruption? E.g. visits to new school, end of pre-school special events<br>What could help with the transition during this time of uncertainty? |

### Analysis and analytical framework

Transcripts were checked for accuracy, anonymised and imported into NVIVO-12 to facilitate data management and analysis. The study team developed an initial coding frame. Three researchers (JC, HT, RB) coded the same three transcripts line-by-line and compared their codes, with inconsistencies discussed and appropriate amendments made to the coding framework. The remaining transcripts were single-coded by these three researchers with ongoing discussion about amending the framework when new codes emerged. These line-by-line codes were then aggregated into broader codes, and later used to develop explanatory themes.

Key concepts from the literature on nurturing proved useful in understanding parents’ accounts and developing our analysis (Dalton, Rapa, & Stein, 2020). Within nurturing frameworks, communication and learning appropriate to children’s developmental stage are essential (Education Scotland & Glasgow City Council, 2017). Understanding that children communicate through their behaviour is also a central component. The importance of routine in helping children to feel safe and secure by providing stability and certainty in their day-to-day lives is recognised too. Finally, these frameworks emphasise the importance of transitions and identify the specific vulnerability of children as they experience normative transitions (such as beginning school) during a period of non-normative transition caused by socio-historical events (i.e. the COVID-19 pandemic). Initial codes were re-grouped around these nurture concepts by SC and developed into three key themes through which we present our data.

## Findings

Participants were predominantly mothers (n=16), White British (n=10), and educated to degree level (n=13), with half the sample living in the highest deprivation quintile (n=10) (see Table 2). Five were single parents. More than half the sample (n=13) reported that at least one parent was not working during the strictest restrictions, with three reporting no household members in employment at the time of interview.

**Table 2.**
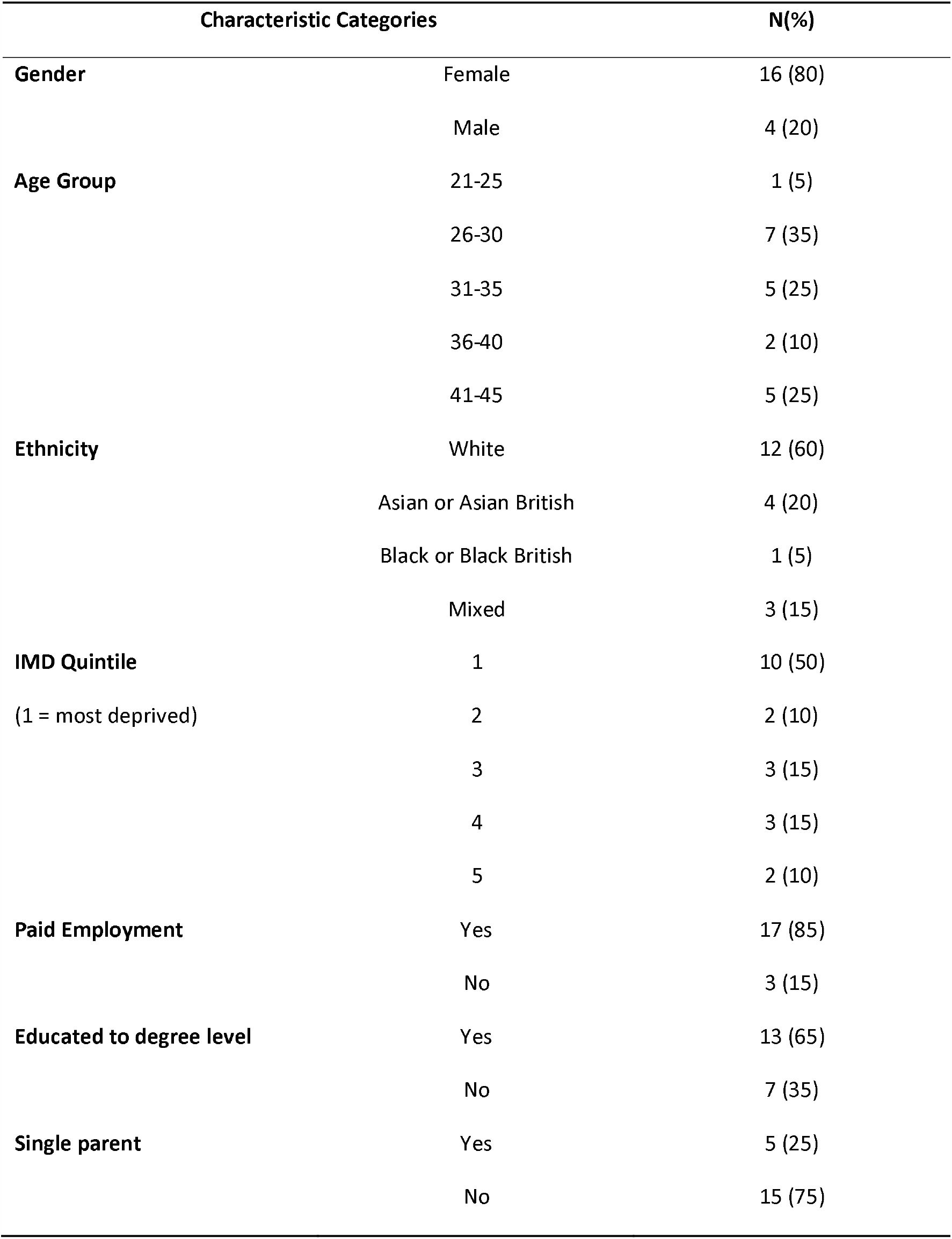
Participant characteristics

### Creating age-appropriate explanations

The sudden onset of COVID-19 restrictions left parents to explain an entirely new and potentially alarming situation to their pre-school child. There were three main areas parents had to manage in terms of supporting their child’s understanding of COVID-19 and restrictions. The first related to what the virus was, transmission and infection, and the hygiene practices necessary to avoid infection. As routines changed and children heard news reports, their anxieties were inevitably raised.

> *He hears things, just through the TV, and especially during the beginning of the pandemic, he was hearing every day how many people had caught the virus and how many people had died from the virus*…*I ended up turning the TV off, so he couldn’t hear it…a child shouldn’t worry about things like that. (Mother_12)*

Parents attempted to explain what was happening in a way that a pre-school child could understand. Parents reported their children had assimilated the new vocabulary (e.g. ‘lockdown’, ‘coronavirus’, ‘germs’) and information about hygiene measures when this information was presented at an appropriate level. They also accepted however that their child’s understanding of the situation was superficial because of their age.

> *With having a four- and a three-year-old, they’re not going to comprehend what COVID-19 is. So we talked about there are germs and we’ve got to stay inside to stay safe, you have to wash your hands when mummy tells you to. It was very difficult for them to process*…*It’s too much for a toddler to comprehend. (Mother_13)*

The second issue parents faced was explaining why certain favoured places, like nurseries and play areas in parks, were no longer accessible. These places were tied to children’s routines and therefore, it was not only that they missed these locations, but also the day-to-day experiences that provided fun and certainty to their lives.

> *I think that’s what he found difficult, is that everything looks the same and everyone seems the same, but he couldn’t do the same things that he normally does. (Mother_6)*

The third issue was ceasing in-person contact with those who were important to their children, such as grandparents and friends. Parents indicated this was one of the most difficult restrictions for children to understand. What had previously been close, nurturing relationships had to be reconceptualised as potentially dangerous and to be avoided.

> *I think it made him really sad, not being able to give his grandma a cuddle. He just couldn’t quite comprehend why he couldn’t do that, even though he knew about the virus and he knew that it can be dangerous, and we’re quite open with the boys and we explained to them how severe it can be for some people. (Mother_6)*

Understanding children’s behaviour Parents reported negative changes in their child’s mood or behaviour during the period of restrictions, including increased tantrums and anger; unsettled, anxious and emotional behaviours; and developmental regression (e.g. sleep, independence). Parents reported children displaying shock, anger, frustration and sadness at the new restrictions placed upon their lives. Parents generally sought to understand why their children were acting in this way, recognising the emotional impact of change in routine, lack of stimulation, not going out as much, reduced social contact and anxiety about coronavirus.

> *He was really emotional. He used to play up a lot. We had no routine, so he had no structure. He was so energetic, I couldn’t burn all his energy off. (Mother_10)*

Although lack of routine was identified by parents as an explanation for changes in children’s behaviour, they found it challenging to create new routines. There were limited opportunities to do this without the support parents previously relied upon, such as childcare, local amenities, family and friends. To reduce the impact of children missing grandparents and others, parents attempted to connect them via technology. Whilst this was positive for a small number of children, for others this was not a means through which they could connect adequately.

> *When we’ve tried to video call some of [my daughter’s] friends or family, she would just get upset, it wasn’t a good enough substitute. (Mother_14)*

Nearly half of parents reported changes in their child’s sleep patterns during restrictions. The main issue reported was difficulty in getting children to fall sleep. Parents often perceived this as an indirect impact of restrictions on their children. Disrupted sleep was viewed as a consequence of reduced physical exertion and increased screen time, but also emotional issues such as boredom and anxiety.

> *We noticed some changes in his sleep pattern, really. Yes, I can say that even in his sleep, sometimes he’s restless. He’s got nightmares. He leaves his bed to come and sleep with us. (Mother_1)*

One parent commented that reduced sleep impacted on her child’s behaviour, indicating the circularity of the challenges families were facing at this time.

> *The longer that [COVID-restrictions have] gone on, like she doesn’t go to sleep at night, and then she’s already irritable before she’s even started the next day. (Mother_5)*

Some parents reported positive changes in their child’s behaviour and mood during the restrictions, which they identified as due to spending more time with the immediate family. These parents reported their children felt secure at home with them, and that this appeared to make them happier.

> *I think she’s been a little bit closer to us because having to spend a little bit more time with us*…*I think in that aspect it’s been good that we have seen her grow and she appreciated the time with us. (Father_2)*

One parent found a positive impact from having more time to set a sleep routine.

> *I work all the time so we hardly got to see each other*…*I was just throwing him into bed at any time*…*Obviously, having lockdown, I had time to do stuff before bedtime. We started having a bath at the same time every night, then story time and then sleep time, which I’ve stuck to. (Mother_10)*

### Concerns about school transition

Parents recognised starting primary school is a major transition for any child, however, they viewed it as a normative transition. It was an expected change in routine that may or may not raise general anxieties for families. Some parents talked about their child seeming ‘ready’ for school. For some there was also a sense of relief that school – and a daily routine – would soon begin.

> *He will start in September, so that will start as a new routine for him… I think we all need it. I think we all need time away from each other. I think it’s not healthy being around each other all the time. (Mother_7)*

Nevertheless, many parents admitted to some concerns about this transition. Some of this was a natural response to their child transitioning to the next stage in life, for example, around toilet training. However, some felt the context of the COVID-19 pandemic made this transition, and their feelings toward it, more complicated. Many normal transition activities (such as in school visits, stay- and-play sessions, contacts with teachers) had been delayed, done ‘virtually’, or cancelled altogether. Some parents reported that schools had been proactive in trying to accommodate transition activities in a way that was COVID-secure, such as through virtual contacts.

> *I think the school have done a really good job. They have sent some videos out that are on YouTube that we have watched which have been really helpful. (Father_2)*

Other parents had not had this experience and were concerned about how they would adequately prepare children for school.

> *I just think the transition is going to be harder because of lockdown, because we haven’t had as much communication with the school as we possibly would have had*…*I think that’s definitely affected the transition for both parents and the child, because if the parents have got no information, they can’t impart anything to the child. (Mother_3)*

Unease due to uncertainty was discussed by many parents. The myriad of unknowns articulated included how to safely drop children at school, where children would eat, the extent to which social distancing would be in place, and whether education would be full-time. Many therefore felt anxiety about what the autumn/winter would bring.

> *How is life going to change for our school child and the circumstances? If another second wave comes then how will life be like in December?*…*Will we be going back to lockdown in December or what will happen during the school time as well? (Mother 11)*

## Discussion

Parents in this study were in no doubt that COVID-19 restrictions had impacted their pre-school children’s emotional wellbeing and they attempted to attend responsively to their children’s needs. They explained the sudden changes age-appropriately and attempted to maintain children’s relationships with extended family and friends within restrictions. Most parents reported negative changes in their child’s behaviour, framing these as understandable responses to lack of routine, boredom, and anxiety. Parents’ options for alleviating these emotions were limited and almost totally reliant on skilfully navigating life at home without external support.

Parents recognised how important starting school was in their children’s development. In the context of the pandemic they were unsure how best to prepare them. Under usual circumstances, they would have a clear understanding of what school life might be like, supported by their nursery and school. Under COVID-19 restrictions, they were instead faced with uncertainty about school life. The potential impact of a lack of sufficient transition has been recognised (Bowyer-Crane et al., 2021). Research with teachers has outlined the barriers in supporting nursery to school transition during COVID-19 restrictions, with skills regression a substantial challenge to be addressed, particularly when working with families from disadvantaged backgrounds and children with special educational needs and disability (Bakopoulou, 2021).

Our findings highlight how parents sought to support and nurture their children through the uncertainty of the pandemic. Masten and Motti-Stefanidi (2020) argue family routines provide safety and comfort to children experiencing severe disruption. A small number of families in our study said they and their children had experienced some positive benefits during the pandemic, such as having an opportunity to bond. A survey of UK parents with young children identified many benefits of spending more time together during the pandemic (Ipsos MORI, 2020). Yoshikawa et al. (2020) hypothesise that increased family closeness may be an important protective outcome of COVID-19 restrictions.

It was clear, however, that families have found it challenging to replace the everyday routines. This may be because parents were expected to do this at a household level. Previous studies of family resilience have stressed the importance of relational approaches where parents are supported by extended family, community-based resources and wider societal structures (Walsh, 2020). During the COVID-19 restrictions, these external relations were largely absent for the parents in this study. Instead, a relational approach focused on their own relationship with their child. Walsh (2020) suggests the best way for parents to support children through ongoing COVID-19 disruption is by creating meaning from challenges, a hopeful outlook and the creation of a sense of purpose. The parents we interviewed were reflecting on a relatively short time frame of only a few months and therefore may not have anticipated that restrictions would continue beyond 2020. In early 2021, restrictions were extended and it is likely that the resilience described by these families was tested, particularly in the winter months, without the wider support of external community resources (Norris, Stevens, Pfefferbaum, Wyche, & Pfefferbaum, 2008). Remote learning was implemented once again for the majority of school-aged children across the UK in January and February 2021, however, in England early years establishments largely remained open. This policy decision perhaps recognised the immense strain families with young children faced in trying to care for them under previous restrictions.

An additional consideration is that our interviews did not ask parents about financial hardship, and none of them raised this spontaneously during the discussions. Half of those interviewed were from households including key workers, who were still earning income. A quarter of households had a member who was furloughed, which may have provided families with a buffer against redundancy, although possibly with their income reduced. For families where parents have lost jobs or have experienced substantial reductions in their income levels, parents’ capacity to manage their children’s emotional wellbeing may have been more limited. As the pandemic and its long-term aftermath continue, a greater proportion of families may be in more perilous positions, leading to increased family stress. Family stress is a particular concern for child welfare during emergency situations, and appears to have had an impact on some families’ experiences during the pandemic. Quantitative surveys of parents have indicated that higher levels of parental mental illness during the pandemic were negatively associated with responsive parenting practices (Brown et al., 2020; Romero, López-Romero, Domínguez-Álvarez, Villar, & Gómez-Fraguela, 2020; Westrupp et al., 2020). A major concern has been that rates of child abuse in England have risen substantially during restrictions. During the first half of 2020 there was an increase of 27% of serious incident notifications and the number of serious incident notifications relating to child deaths increased by 30 to 119 compared with the same period in the previous year, with the largest increases seen in young children.

### Strengths and limitations

This study offers novel and in-depth insights into how parents with a pre-school child managed their children’s emotional wellbeing in response to COVID-19 restrictions. It complements quantitative surveys (Dodd et al., 2020; Public Health Scotland, 2020) by providing in-depth understanding of the stresses parents and their children face. These findings will be important for improving the support provided to families during future restrictions and informing plans for longer-term recovery (Hefferon et al., 2020). We note however that we only include the perspectives of parents, rather than the children themselves.

Using a nurturing lens helped to develop the themes identified at an earlier analysis stage and provided interpretative insight into early child development. The results indicated that parents were engaging in positive parenting practices that could be described as responsive and nurturing. We acknowledge, however, that parents who were struggling to care for their children during COVID-19 restrictions might be less likely to participate in a study such as this, or to disclose less optimal parenting strategies in the interview context.

The study had strong representation of families living in areas of high deprivation and of different ethnicities, providing the opportunity to ensure multiple perspectives were considered. Nevertheless, the sample was highly educated, and may not reflect the experiences of parents with lower levels of education. Another study limitation was that it only focused on one aspect of family life at this time, specifically parents’ experiences of their pre-school child’s emotional wellbeing. There was only limited time within the interview to discuss wider family life, including parents’ emotional wellbeing and wider family relationships, which are also likely to be impacting on young children’s emotional wellbeing.

### Implications and conclusion

This study is one of the first to provide an in-depth understanding of parental perceptions of young children’s emotional wellbeing under COVID-19 restrictions. With that in mind, it has relevance to families and the professionals who support families globally. Although young children may be unable to understand in substantial detail what the virus is, they undoubtedly experience the monumental disruption it brings to their young lives. Routines that brought them comfort and security were disrupted overnight, with only their parents available to support them.

The long-term impacts of this disruption so early in life are not yet understood, but studies examining families’ experiences at this time will provide a foundation to build appropriate support for the future. The nurturing framework used in this study has highlighted the importance of creating secure environments for children to thrive in. Nurture is a possible mitigation strategy to reduce the negative impact of the pandemic on children’s wellbeing. It can also be extended to form the basis of recovery too, including a coordinated response from services to support families to create strong, positive relationships and understand behaviour as communicating important information about a child’s emotional wellbeing. Suggested responses thus far have included extending the school day, or reducing the length of school holidays to allow children to catch up on lost learning. These recovery options do not acknowledge the need for children’s wellbeing to be addressed first, as is embedded in nurture approaches in educational settings, before learning can take place optimally. We therefore propose that services and communities work to nurture children and their families’ wellbeing. This will help parents to offer a complementary responsive approach to their children as they support them through and beyond the COVID-19 pandemic.

## Data Availability

Anonymised study data will be made available under controlled access via the University of Bristol data repository. Requests for controlled data will be referred to an appropriate Data Access Committee for approval before data can be shared with bona fide researchers, after their host institution has signed a Data Access Agreement.

https://www.bristol.ac.uk/staff/researchers/data/accessing-research-data/

## Acknowledgements

We are very grateful to all the parents who took part in the research interviews and to the nurseries who helped support recruitment. The views expressed in this publication are those of the authors and not necessarily any of the funding bodies listed.

